# Automated Task-Specific vs General-Purpose Artificial Intelligence for Detecting Subtle Intraoperative Warning Signs During Cataract Surgery: A Multicenter Diagnostic Study

**DOI:** 10.64898/2026.01.15.26344200

**Authors:** Ye Zhang, Lei Chen, Wenhao Zhao, Hui Zhang, Chunyan Qiao, Zijian Liu, Charlotte Hui Chung, Marcus Chun Jin Tan, Meng Wang, Yih Chung Tham, Victor Koh, Chingyu Cheng, Dianbo Liu

**Affiliations:** Yong Loo Lin School of Medicine, National University of Singapore, Singapore; Beijing Tongren Eye Center, Beijing Key Laboratory of Ophthalmology and Visual Science, Beijing Tongren Hospital, Capital Medical University, Beijing, China; Department of Public Health and AI, National Cancer Center Graduate School of Cancer Science and Policy, Korea; Department of Biomedical Engineering and Department of Electrical and Computer Engineering, National University of Singapore, Singapore; Singapore Eye Research Institute, Singapore National Eye Centre, Singapore; Eye Academic Clinical Program, Duke NUS Medical School, Singapore; Ophthalmology & Visual Sciences Academic Clinical Program (EYE ACP), Duke-NUS Medical School, Singapore, Singapore

## Abstract

**Importance:** Early intraoperative warning signs of zonular instability during cataract surgery, such as anterior capsular radial folds, are subtle and easily missed but are clinically important for preventing surgical complications. Whether current artificial intelligence (AI) systems can reliably detect such subtle warning signs in real-world surgical video remains unknown. Recently, automated AI model generators have become available, enabling the automatic construction of task-specific AI models for individual clinical tasks.

**Objective:** To evaluate the diagnostic performance of general-purpose and automated task-specific artificial intelligence systems for detecting anterior capsular radial folds during cataract surgery and to compare their performance with human clinicians.

**Design, Setting, and Participants:** This retrospective diagnostic study used 537 continuous curvilinear capsulorhexis (CCC) video clips collected from Beijing Tongren Hospital (China), National University Hospital (Singapore), and the OphNet-APTOS public dataset.

**Exposure:** Presence or absence of anterior capsular radial folds during CCC, annotated at both clip and frame levels by senior glaucoma surgeons based on expert consensus.

**Main Outcomes and Measures:** Discrimination between fold-positive and fold-negative cases was assessed using macro-averaged precision, recall, and F1 score at the frame and clip levels. Performance was compared among general-purpose AI systems, task-specific models generated by an automated AI model generator, and human graders with different levels of clinical experience.

**Results:** Among 537 video clips (mean 7.32 seconds), 156 (29.1%) were fold-positive. General-purpose AI systems showed limited and inconsistent performance; the best-performing model achieved a mean F1 score of 0.519, and fine-tuned models remained inferior to human graders (maximum F1 score, 0.606). In contrast, task-specific models generated by an automated AI model generator achieved substantially higher performance (F1 score, 0.869; area under the receiver operating characteristic curve, 0.958). In head-to-head comparison with clinicians, the top automated task-specific model (F1 score, 0.835) matched the performance of junior specialists (mean F1 score, 0.829) but remained below that of senior specialists.

**Conclusions and Relevance:** General-purpose artificial intelligence systems do not reliably detect subtle intraoperative warning signs during cataract surgery and consistently underperform human clinicians. In contrast, recently available automated AI model generators enable the creation of task-specific models with near–junior specialist performance. These findings suggest that clinically reliable surgical AI is more likely to be achieved through automated generation of task-specific models rather than through general-purpose AI systems. Although evaluated in cataract surgery, these findings highlight a broader challenge for artificial intelligence in detecting brief, low-contrast intraoperative warning signs in surgical video.

**Key Points:** *Question:* How reliably can general-purpose artificial intelligence (AI) systems and task-specific AI models generated by an automated AI model generator detect subtle intraoperative warning signs during cataract surgery compared with human clinicians?

*Findings:* In this multicenter diagnostic study of 537 cataract surgery video clips, general-purpose AI systems were unreliable and consistently underperformed human clinicians in detecting anterior capsular radial folds. In contrast, task-specific AI models generated by an automated AI model generator—a technology that has only recently become available—achieved substantially higher diagnostic performance and matched the performance of junior specialists.

*Meaning:* General-purpose AI systems show limited reliability for detecting subtle intraoperative warning signs during cataract surgery. The recent availability of automated AI model generators enables a new paradigm of task-specific model development and represents a more clinically viable path for surgical decision support.

## Introduction

General-purpose artificial intelligence (AI) systems, including vision–language models (VLMs), have demonstrated rapid progress in aligning visual and textual data, driving interest in “generalist” AI systems designed to handle diverse tasks using a single pre-trained backbone. However, these models rely on broad training data that may lack the granular specificity required for high-stakes surgical diagnostics.¹⁻³ Even when fine-tuned, general-purpose AI systems remain constrained by their underlying architectures and often struggle to adapt to the unique visual domains of specific medical problems. Consequently, existing benchmarks, which predominantly focus on static medical images or curated visual question–answering tasks, do not evaluate whether a single general-purpose AI system can reliably resolve the subtle, transient, and low-contrast cues that characterize real-world operative video, or whether their broad training introduces noise that obscures fine-grained clinical signals.⁴⁻⁶

In parallel, a new paradigm has recently emerged in which automated AI model generators are used to construct task-specific models tailored to individual clinical problems. Rather than relying on direct inference from a fixed general-purpose model, these systems automatically generate executable code to build, train, and optimize bespoke supervised classifiers for a given task. By constructing a dedicated solution optimized for the specific constraints of a dataset, automated task-specific model generation may offer a more robust and clinically reliable approach to medical image analysis than one-size-fits-all general-purpose AI systems.⁷˒⁸ However, whether this task-specific, automated approach offers superior diagnostic utility for complex surgical video interpretation remains unverified.

To address this uncertainty, we evaluated these two strategies using anterior capsular radial folds at the initiation of continuous curvilinear capsulorhexis (CCC),a subtle but clinically important intraoperative sign suggestive of zonular instability during cataract surgery,as a stringent test case for fine-grained visual discrimination.⁹˒¹⁰ Cataract surgery provides a stringent test case for surgical artificial intelligence: visual cues are subtle, transient, and embedded within high-motion operative video. Failure to detect these cues can lead to downstream complications, making this domain a useful lens for examining the clinical reliability of current AI paradigms. Prior work in ophthalmic AI has addressed surgical instrument detection, phase recognition, and risk prediction, but existing benchmarks have not systematically evaluated subtle intraoperative warning signs or compared general-purpose AI systems with automated task-specific model generation.¹¹⁻¹⁵ We curated a multicenter dataset of annotated capsulorhexis video clips and compared the diagnostic performance of leading general-purpose AI systems with task-specific models generated by an automated AI model generator. Furthermore, we contextualized these findings against human graders with varying levels of clinical experience to assess whether automated task-specific modeling can bridge the performance gap between current AI systems and clinical expertise.

## Methods

### Dataset, Annotation and Task

A multi-source dataset was assembled from three independent cohorts, restricted to the CCC phase. This phase was selected because anterior capsular radial folds typically manifest at CCC initiation, when the capsule is punctured and traction is transmitted through zonular fibers.

#### Beijing Cohort

A total of 215 phacoemulsification videos were retrospectively collected from Beijing Tongren Hospital under Institutional Review Board (IRB) approval. After excluding clips with inadequate visualization, 201 videos were included. Among these, 94 (46.8%) demonstrated radial folds, while 107 (53.2%) did not. Demographic characteristics are shown in Table 1.

**Table 1.**
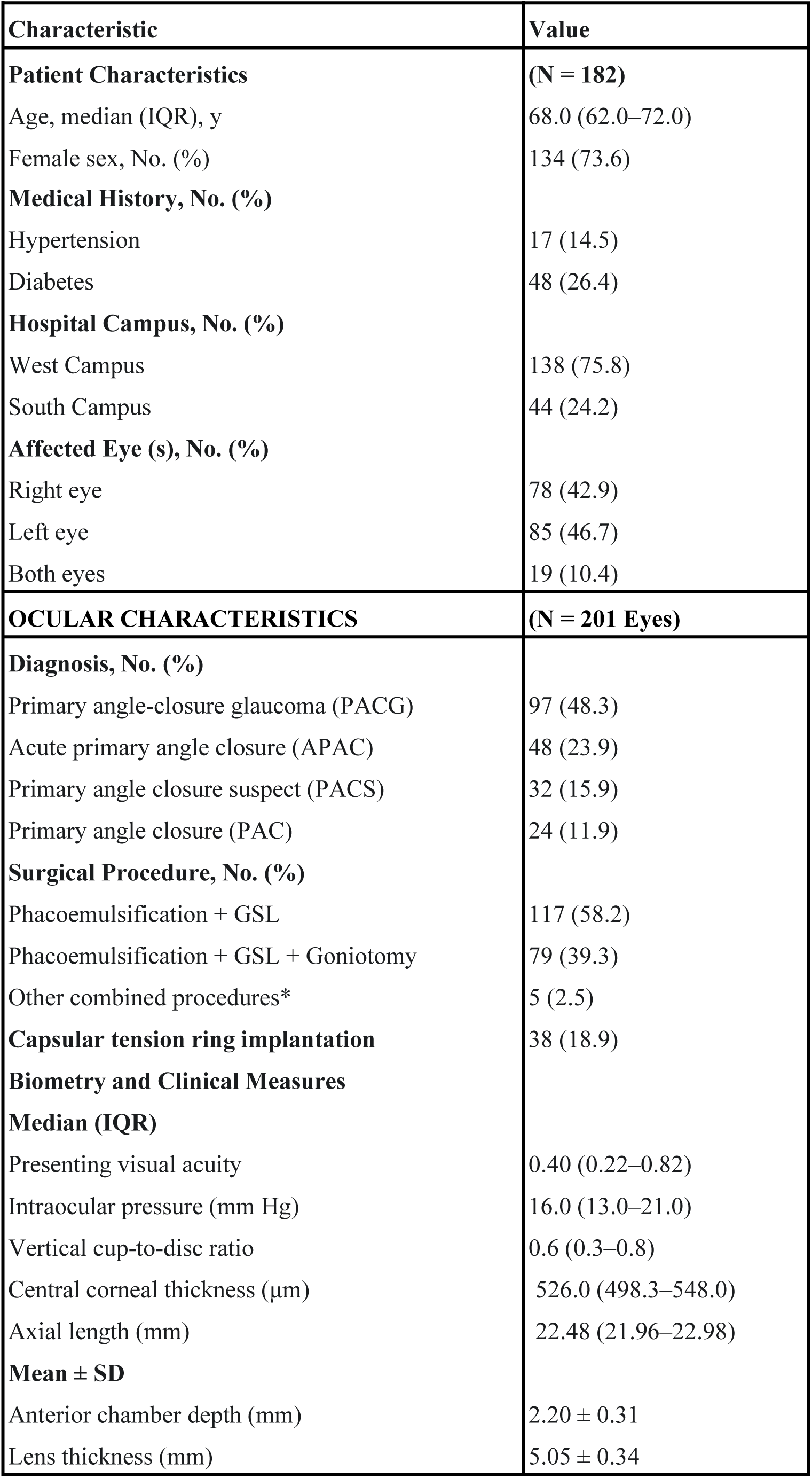
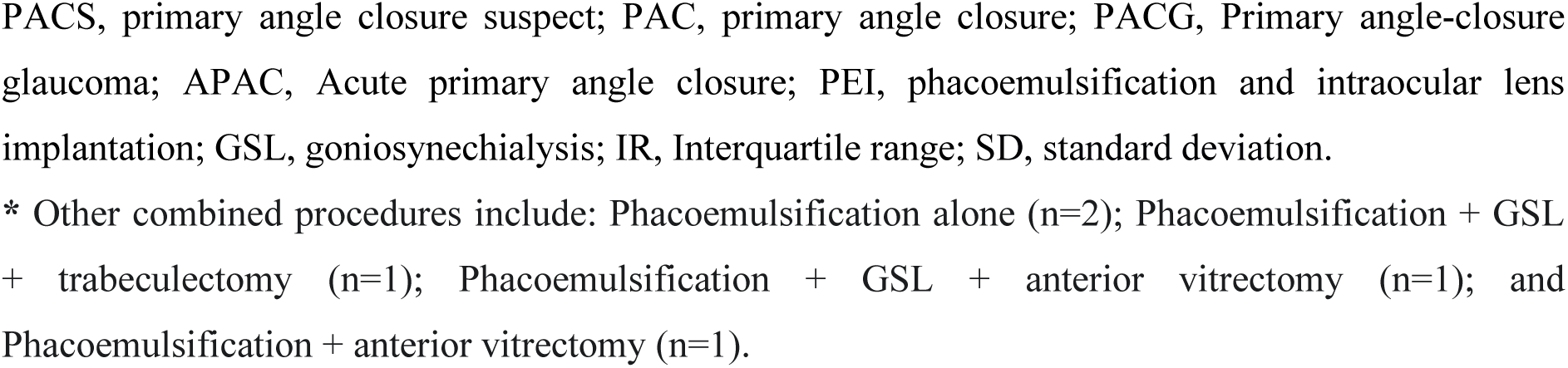
Demographic and Ocular Characteristics of Patients from Beijing Cohort

| Characteristic | Value |
| --- | --- |
| **Patient Characteristics** | **(N = 182)** |
| Age, median (IQR), y | 68.0 (62.0–72.0) |
| Female sex, No. (%) | 134 (73.6) |
| **Medical History, No. (%)** |  |
| Hypertension | 17 (14.5) |
| Diabetes | 48 (26.4) |
| **Hospital Campus, No. (%)** |  |
| West Campus | 138 (75.8) |
| South Campus | 44 (24.2) |
| **Affected Eye (s), No. (%)** |  |
| Right eye | 78 (42.9) |
| Left eye | 85 (46.7) |
| Both eyes | 19 (10.4) |
| **OCULAR CHARACTERISTICS** | **(N = 201 Eyes)** |
| **Diagnosis, No. (%)** |  |
| Primary angle-closure glaucoma (PACG) | 97 (48.3) |
| Acute primary angle closure (APAC) | 48 (23.9) |
| Primary angle closure suspect (PACS) | 32 (15.9) |
| Primary angle closure (PAC) | 24 (11.9) |
| **Surgical Procedure, No. (%)** |  |
| Phacoemulsification + GSL | 117 (58.2) |
| Phacoemulsification + GSL + Goniotomy | 79 (39.3) |
| Other combined procedures* | 5 (2.5) |
| **Capsular tension ring implantation** | 38 (18.9) |
| **Biometry and Clinical Measures** |  |
| **Median (IQR)** |  |
| Presenting visual acuity | 0.40 (0.22–0.82) |
| Intraocular pressure (mm Hg) | 16.0 (13.0–21.0) |
| Vertical cup-to-disc ratio | 0.6 (0.3–0.8) |
| Central corneal thickness (μm) | 526.0 (498.3–548.0) |
| Axial length (mm) | 22.48 (21.96–22.98) |
| **Mean ± SD** |  |
| Anterior chamber depth (mm) | 2.20 ± 0.31 |
| Lens thickness (mm) | 5.05 ± 0.34 |

PACS, primary angle closure suspect; PAC, primary angle closure; PACG, Primary angle-closure glaucoma; APAC, Acute primary angle closure; PEI, phacoemulsification and intraocular lens implantation; GSL, goniosynechialysis; IR, Interquartile range; SD, standard deviation.
\* Other combined procedures include: Phacoemulsification alone (n=2); Phacoemulsification + GSL + trabeculectomy (n=1); Phacoemulsification + GSL + anterior vitrectomy (n=1); and Phacoemulsification + anterior vitrectomy (n=1).

#### Singapore Cohort

Thirty videos were obtained from National University Hospital, Singapore. Following screening for completeness, 27 were included for analysis. Of these, 6 (22.2%) demonstrated radial folds and 21 (77.8%) showed no folds.

#### OphNet–APTOS Public Dataset

The OphNet–APTOS repository was reviewed to supplement external data.^16^ From 496 cataract surgery videos, 308 evaluable CCC clips were selected (one video contained two distinct segments). Expert annotation identified 56 (18.1%) clips with visible radial folds and 253 (81.9%) without.

#### Final Dataset

The combined dataset comprised 537 clips, with 156 (29.1%) labeled as fold-positive and 381 (70.9%) as fold-negative. All clips were limited to the CCC phase, with durations ranging from 5 to 20 seconds (mean ± standard deviation [SD]: 7.32 ± 3.35 s).

Labeling was performed in two stages. Clip-level labels were assigned by two experienced glaucoma surgeons via consensus based on intraoperative observation (Beijing) or video review (Singapore/OphNet). Clips were classified as fold-positive or fold-negative based on visible radial folds at CCC initiation. Frame-level annotation was performed independently by a single experienced glaucoma surgeon. Videos were sampled at 1–2 fps and each frame underwent a two-step review: (1) identification of active tearing initiation (instrument contact with traction), and (2) assessment of radial fold presence. Frames representing pauses were excluded, resulting in a dataset of discrete tearing images.

Automated detection was formulated as a binary classification task (fold-positive vs. fold-negative). To evaluate diagnostic capabilities, we defined three distinct levels of analysis, ranging from static image recognition to temporal event understanding. Frame-Level Analysis: Evaluates perceptual limits using single static frames, isolating visual feature detection without temporal context. Clip-Level Analysis: Simulates a screening workflow favoring sensitivity. Using an “OR” aggregation rule, a clip is classified as positive if the model detects pathology in any constituent frame. Frame-Sequence Analysis: Assesses contextual reasoning using continuous video clips. The model processes the temporal sequence to distinguish pathological events from motion artifacts.

Two complementary task settings were established to test these levels: Vision-Language models Inference, where models predicted folds in zero-shot or few-shot modes using frames or sequences (Supplementary A); and Automated Generator-driven Classification, where AI model generators were provided only with image paths and labels to generate executable training pipelines. Due to the complexity of temporal architectures, the output of these generators was restricted to frame-level classification.

### General-Purpose Visual language Models Detection and Evaluation

A representative set of widely used general-purpose and medical-specific vision–language models was evaluated in this study to assess whether model generality, rather than a specific architecture, limits detection of subtle intraoperative cues. We evaluated 11 Vision-Language Models utilizing their publicly released checkpoints without fine-tuning. These included 2 proprietary models (GPT-4.1, Gemini 2.5 Pro) and 9 open-source general-purpose or medical-specific models included Qwen2.5-VL (7B)^17^ and InternVL-3.5 (8B).^18^ Medical-specific models included Lingshu (7B),^19^ HuatuoGPT-Vision (7B),^20^ MedGemma (4B),^21^ HealthGPT,^22^ QoQ-Med-VL (7B),^23^ LLaVA-Med (v1.5-7B),^24^ and the ophthalmology-focused OphthaReason.^25^

Inference was conducted at a uniform sampling rate of 2 frames per second. For single-frame analysis, static images were evaluated using zero-shot text prompts. Three prompt variants were tested (Supplementary B). For models supporting multiple inputs, a few-shot condition was added using three reference images (one normal case, two abnormal cases [mild and severe]) to aid in resolving subtle deformations. Additionally, continuous frame sequences were processed simultaneously to leverage temporal context for holistic prediction (Supplementary C). Performance was assessed using macro-averaged precision, recall, and F1-score across three levels of granularity: frame-level; clip-level; and frame-sequence level.

### Automatic AI Model Generators Detection and Evaluation

Four automated AI model generators (CodeX, Qoder, AIDE,^26^ and GitHub Copilot powered by Claude Sonnet 4.5) with generating executable code for supervised image classification. The dataset was structured to ensure rigorous evaluation: Beijing and OphNet-APTOS cohorts were combined and split into training (60%), validation (20%), and internal test (20%) sets, while the Singapore cohort as an external test set. Guided by a standardized prompt (Supplementary D) and input CSV files, the generators executed an autonomous two-stage workflow. This process progressed from an initial code generation phase to an iterative optimization stage, where generators analyzed execution logs, error reports, and preliminary results to implement targeted refinements, such as modifying loss function weighting or tuning prediction thresholds. The final outputs were evaluated based on classification performance and code quality, utilizing an LLM-as-judge framework to assess modularity, documentation, robustness, engineering best practices, and efficiency (Supplementary E).

### Human–AI Frame-Level Comparison

To contextualize AI performance, a single-frame classification benchmark was conducted using 729 frames (224 fold-positive) extracted from 100 expert-validated clips (50 positive, 50 negative). This setting isolated visual pattern recognition to ensure consistency with frame-level AI inference.

Seven human graders participated, stratified by experience: six glaucoma specialists from Beijing Tongren Hospital (two junior, two intermediate, two senior) and one medical student from Singapore. Graders independently classified frames while masked to ground truth, class distribution, and AI predictions. Standard metrics (macro-averaged) were calculated for individual graders and summarized by experience level to provide a comparative baseline.

## Results

### Performance of General Purpose AI Models

Performance was evaluated across frame, clip, and frame-sequence levels using macro-averaged metrics (Supplementary F). At the frame level, GPT-4.1 benefited substantially from few-shot visual context, with F1-scores rising from 0.177 ± 0.035 (zero-shot) to 0.480 ± 0.055 (Figure 2[a]). Gemini 2.5 Pro exhibited moderate zero-shot performance (F1: 0.307 ± 0.134) with high sensitivity to prompt variations. Among open-source models, Qwen2.5-VL achieved the strongest overall results (few-shot F1: 0.519 ± 0.029). Notably, the medical-specific QoQ-Med-VL demonstrated superior stability, achieving a few-shot F1-score of 0.485 ± 0.004 and outperforming general-purpose models like InternVL-3.5 in zero-shot settings. Conversely, the ophthalmology-focused OphthaReason displayed inverse scaling, performing significantly better with text-only instructions (F1: 0.386 ± 0.155) than with visual examples (F1: 0.245 ± 0.172). Other medical models, including Lingshu, MedGemma, and LLaVA-Med, showed limited discriminative capacity, with F1-scores often falling below 0.170. HuatuoGPT-Vision and HealthGPT showed marked variability, ranging from functional performance to near-failure depending on prompt formulation.

**Figure 1.**
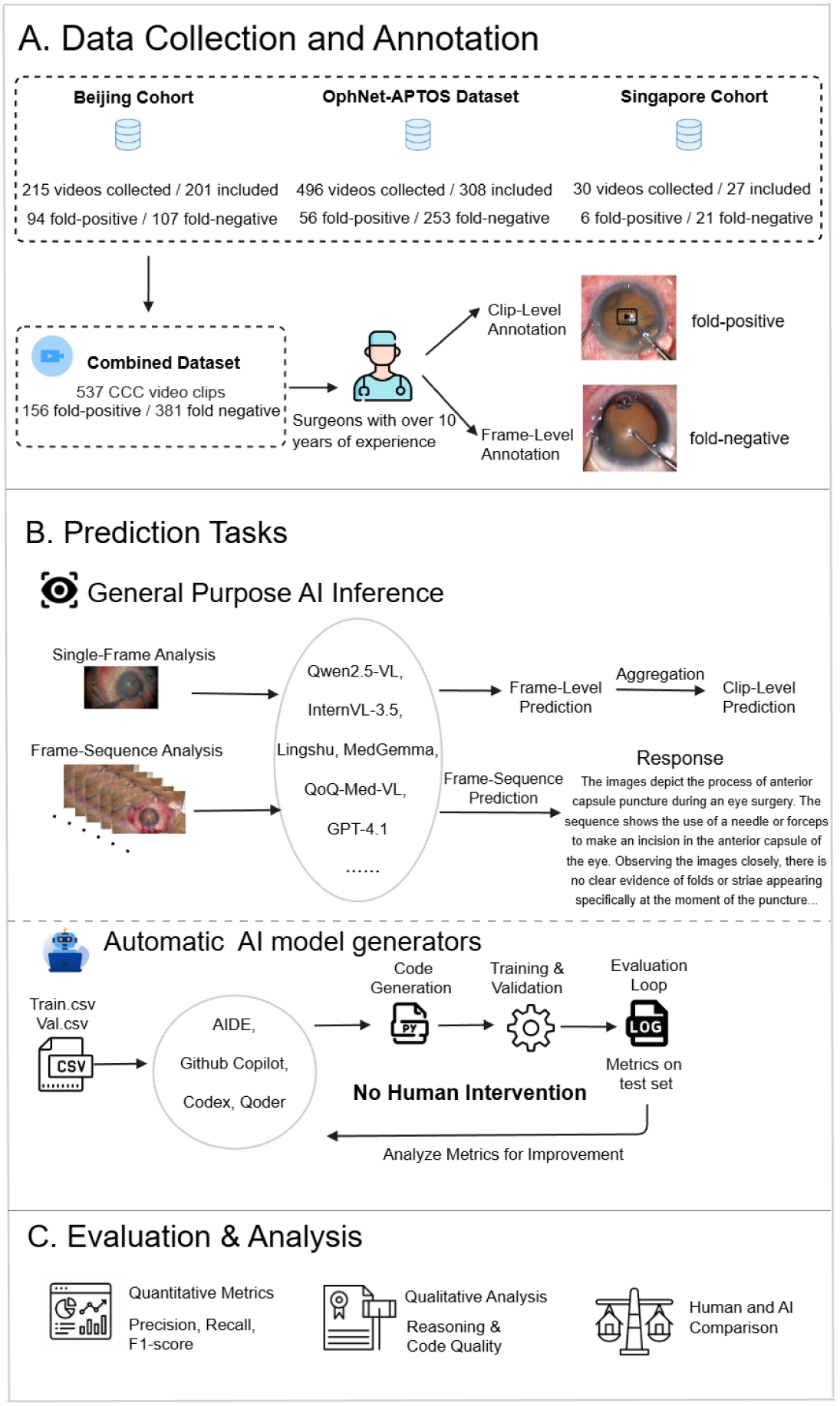
Overview of the study workflow. **(A) Data Collection and Annotation:** A multi-center dataset of 537 continuous curvilinear capsulorhexis video clips were curated from three cohorts (Beijing, OphNet-APTOS, and Singapore) and annotated by experienced surgeons at both clip and frame levels. **(B) Prediction Tasks:** The study evaluates two AI paradigms: (1) **General-purpose AI systems (Vision-Language Models) Inference**, where models perform direct reasoning on single frames (aggregated to clip level) or frame sequences via zero-shot and few-shot; and (2) **Automatic AI model generators**, which autonomously generate, debug, and optimize code to train supervised classifiers using pre-partitioned CSVs. **(C) Evaluation & Analysis:** Comprehensive assessment includes quantitative metrics, a qualitative analysis of both the reasoning patterns of general-purpose AI and the quality of code produced by the automatic generators, alongside a comparison against human experts.

**Figure 2.**
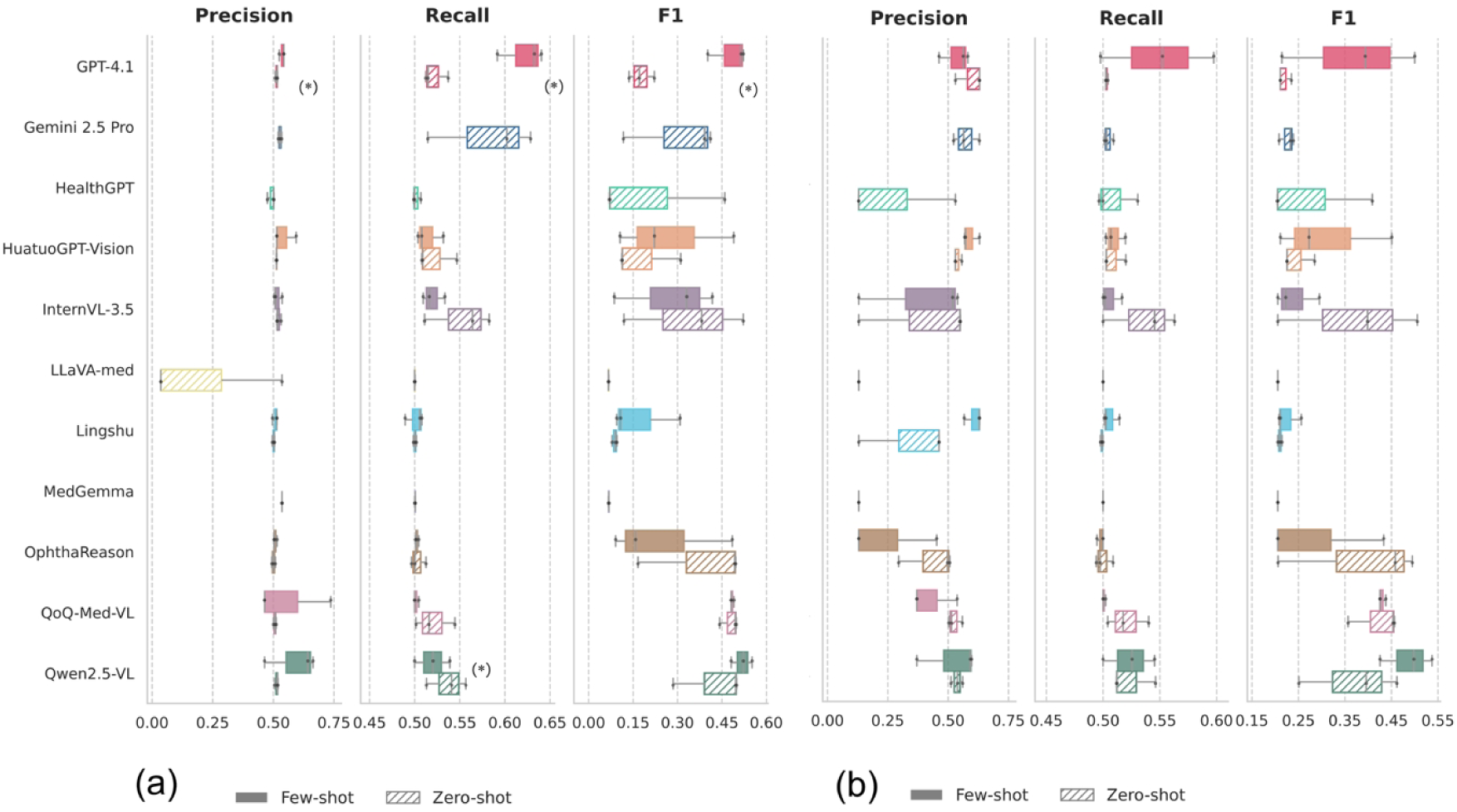
Performance of Vision-Language Models in Detecting Anterior Capsular Radial Folds. Comparison of model performance metrics (Precision, Recall, and F1-score) at the frame level (a) and aggregated clip level (b) via single-frame analysis. Models were evaluated under zero-shot (striped bars) and few-shot (solid bars) inference settings. Data are presented as the mean of 3 prompt variations; error bars indicate the Standard Deviation (SD). In the frame-level analysis, few-shot prompting yielded statistically significant improvements for GPT-4.1 and Qwen2.5-VL compared with zero-shot inference. *P < .05.

Clip-level aggregation amplified these trends (Figure 2[b]). Qwen2.5-VL remained the top performer (few-shot F1: 0.487 ± 0.046). GPT-4.1 improved with few-shot inputs (F1: 0.369 ± 0.118) but exhibited constrained recall in zero-shot modes. QoQ-Med-VL maintained high stability (F1: 0.430 ± 0.006), again surpassing general-purpose models in zero-shot tasks. OphthaReason continued its negative trend with few-shot prompting, while MedGemma and LLaVA-Med produced degenerate predictions with no variance. Frame-sequence analysis yielded similar hierarchies (Supplementary E). Qwen2.5-VL achieved the highest mean F1-score (0.461 ± 0.054) but with significant precision variability. QoQ-Med-VL followed closely (F1: 0.432 ± 0.011) with reduced volatility across prompting strategies. InternVL-3.5 and HuatuoGPT-Vision showed moderate performance but struggled to balance precision and recall, while Lingshu demonstrated limited effectiveness for temporal analysis.

Qualitative analysis of textual rationales and spatial attention provided further insights into performance variability. Recurrent errors included the misinterpretation of surgical stages, frequently confusing initial punctures with later capsulorhexis phases, and the failure to recognize subtle, transient deformations. Classification was further hindered by temporal confounders, such as specular reflections. Moreover, spatial attention analysis using relative attention normalization,^27^ which computes layer-wise attention ratios between task-specific and generic prompts from the final prompt token during the prefill phase, revealed consistently diffuse and poorly localized activation maps (Figure 3). Across these models, attention was predominantly distributed toward surgical instruments, suture lines, and periocular tissue rather than concentrated at the capsular puncture site itself. Even higher-performing models exhibited broad attention across the surgical field, indicating reliance on global contextual cues rather than anatomically focused visual grounding of the subtle capsular deformations critical for accurate classification.

**Figure 3.**
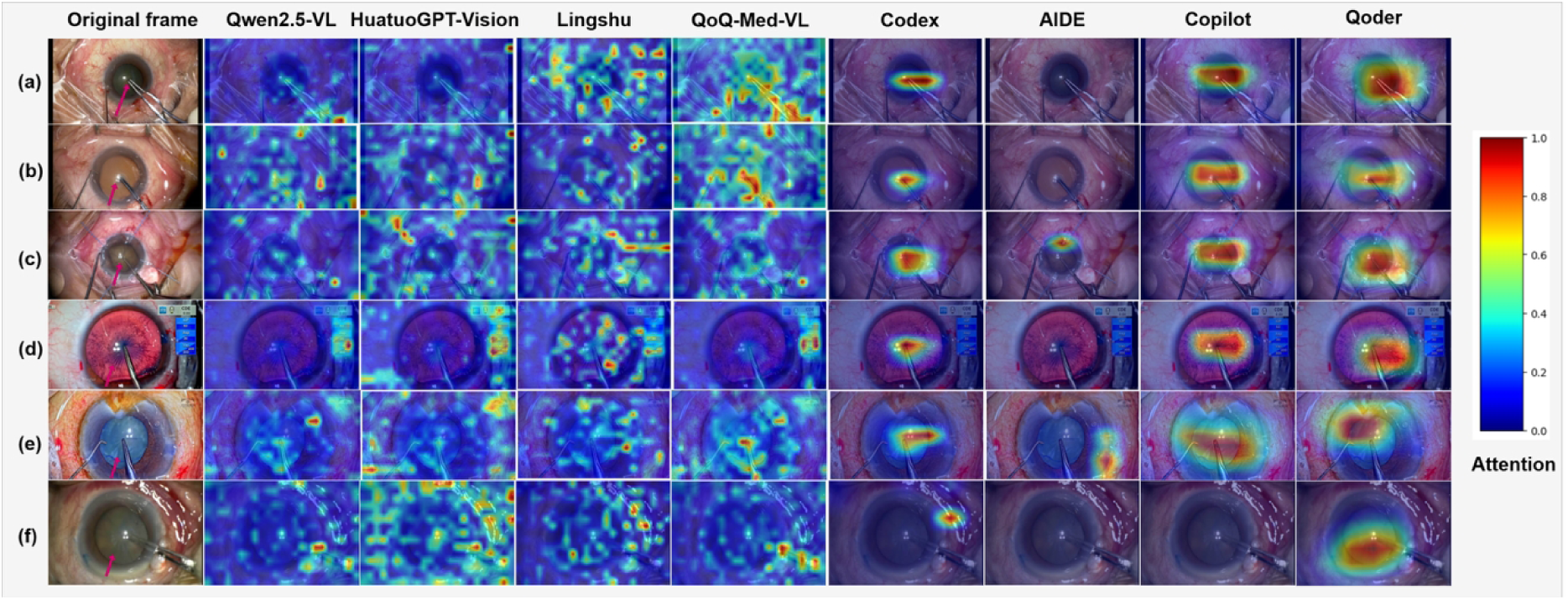
Illustrative Attention Patterns in General-Purpose and Task-Specific AI Models. Representative examples illustrating how different artificial intelligence (AI) approaches attend to visual information during detection of anterior capsular radial folds in cataract surgery. Column 1 shows the original surgical frames, with arrows indicating the expert-annotated location of the intraoperative finding. Columns 2–5 display attention maps from general-purpose vision–language models, and columns 6–9 display activation maps from task-specific supervised classifiers generated by automated AI model generators. Across cases (rows a–f), general-purpose models demonstrate diffuse attention across the surgical field, whereas task-specific models show more localized activation centered on the capsulorhexis region. Warmer colors indicate regions of higher model activation. Details of attention map generation are provided in the Methods.

### Performance of automatic AI model generators

Quantitative evaluation of the frame-level classifiers synthesized by these generators revealed distinct performance tiers (Supplementary G). GitHub Copilot achieved the highest overall effectiveness with an F1-score of 0.869 and a robust area under the receiver operating characteristic curve (AUROC) of 0.958. Qoder demonstrated the highest discrimination capability (AUROC: 0.960, F1: 0.859), while CodeX delivered comparable strong results (F1: 0.850, AUROC: 0.954). In sharp contrast, AIDE failed to produce a viable classifier, suffering from mode collapse (F1: 0.484, AUROC: 0.500) and an inability to differentiate between fold-positive and fold-negative frames.

Grad-CAM visualizations (Figure 3) corroborated these findings^28^. Models generated by the top-performing generators (Copilot, Qoder, and CodeX) exhibited localized attention centered precisely on the capsulorhexis region, aligning with their superior quantitative metrics. Conversely, AIDE-generated models displayed diffuse, scattered attention patterns, failing to converge on anatomically relevant structures.

### Human and AI Comparison Results

To contextualize AI performance, frame-level classification was compared against seven human graders of varying experience using a held-out set of 100 clips. Human performance generally correlated with clinical seniority. Senior glaucoma specialists achieved the highest mean F1-score (0.905), characterized by balanced precision and recall. Intermediate specialists exhibited higher variability (mean F1: 0.810), while junior residents (mean F1: 0.829) and the medical student (F1: 0.830) showed consistent, comparable performance.

Benchmarking revealed distinct performance stratification between model types (Table 2). Vision-Language Models consistently underperformed all human graders. The best proprietary model, GPT-4.1, achieved an F1-score of 0.587, indicating a substantial gap in domain-specific recognition. Similarly, Qwen2.5-VL remained inferior to novice human graders even after fine-tuning (F1: 0.606), underscoring the limitations of generalist multimodal reasoning for detecting subtle intraoperative cues.

**Table 2.**
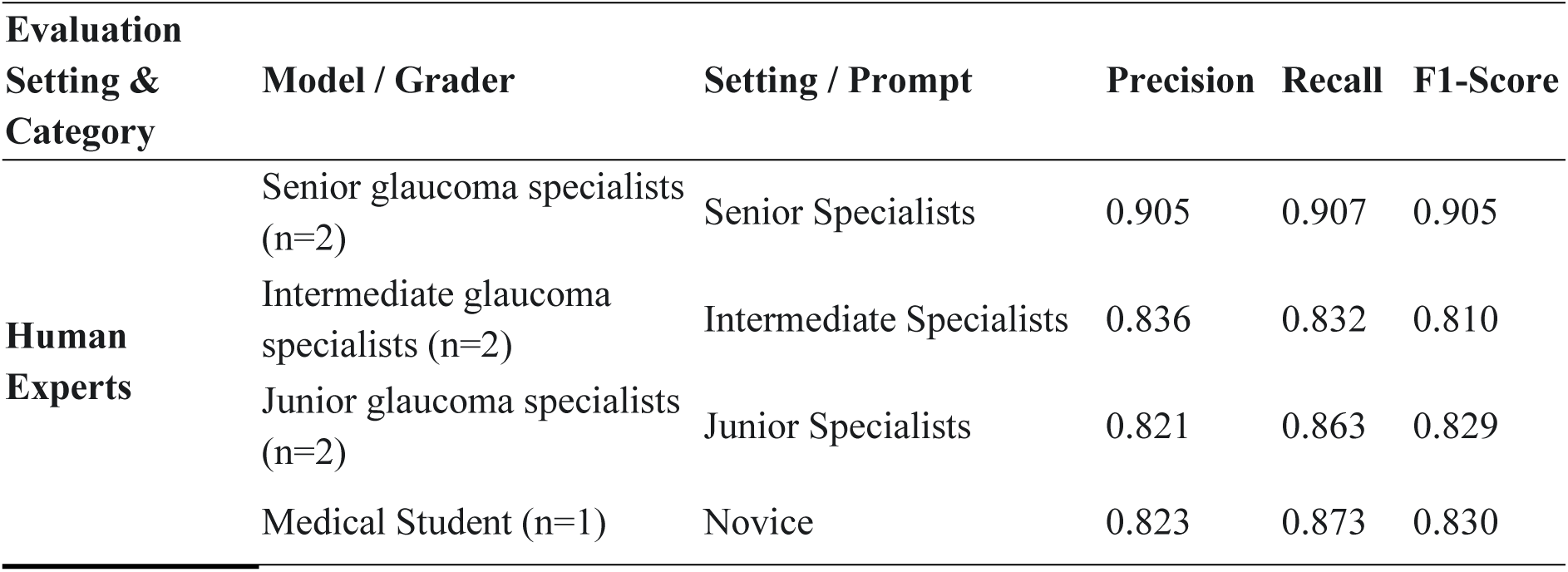
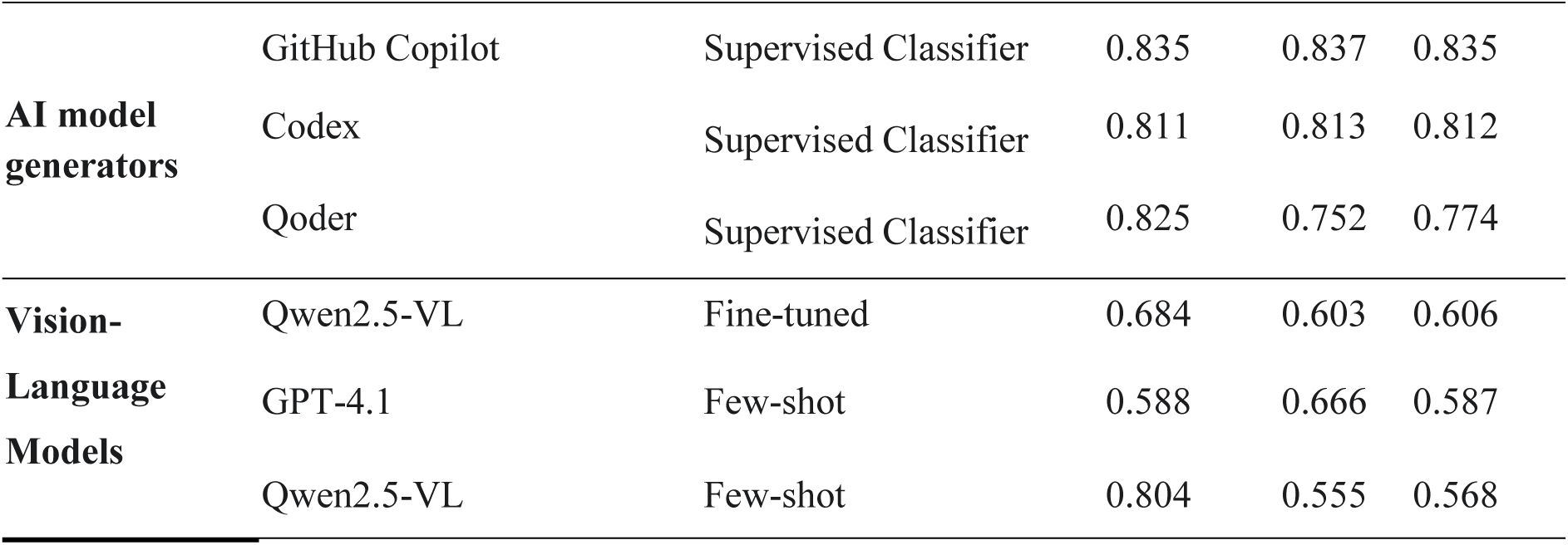
Comprehensive Performance Comparison: Humans, automated AI model generators, and Vision-Language Models. This table presents a direct comparison of the top-performing AI model generator and Vision-Language models against human graders on a held-out subset of 100 video clips. All classification metrics are macro-averaged. Data are presented as Mean ± Standard Deviation for each metric and setting.

In contrast, the supervised classifiers produced by automated AI model generators approached human-level competence. GitHub Copilot achieved a macro-averaged F1-score of 0.835, matching junior specialists (0.829) and the medical student (0.830), and surpassing the intermediate group average. CodeX also demonstrated competitive performance (F1: 0.812), comparable to attending physicians. While Qoder trailed slightly (F1: 0.774) due to lower recall, all top-tier automated generators significantly outperformed Vision-Language models. Although no generator reached the expert ceiling established by senior specialists (mean F1: 0.905), these results demonstrate that autonomously constructed, task-specific classifiers can operate at a clinically meaningful level.

## Discussion

This study evaluated general-purpose artificial intelligence (AI) systems and automated task-specific AI model generators on their ability to resolve brief, low-contrast cues in surgical video, using anterior capsular radial folds during continuous curvilinear capsulorhexis (CCC) as a constrained test case. Across 537 annotated multicenter video clips, we compared zero-shot and few-shot inference by general-purpose AI systems with task-specific models generated by an automated AI model generator and contextualized model performance against human graders with varying levels of clinical experience. These findings highlight both the potential and current limitations of general-purpose AI for fine-grained surgical video interpretation and the promise of automated task-specific modeling for clinically reliable deployment.

### Principal Findings

This work introduces a multicenter surgical video dataset focused on phacoemulsification, providing a reproducible framework for evaluating fine-grained intraoperative classification under real-world variability. By integrating data from two Asian surgical centers (Beijing and Singapore) and a publicly available international repository, the dataset captures heterogeneity in surgical technique, imaging conditions, and operative context that is essential for benchmarking algorithmic generalization.

Evaluation of general-purpose AI systems revealed a landscape characterized by strong prompt sensitivity and architectural dependence. Few-shot prompting partially mitigated performance deficits—for example, increasing GPT-4.1’s F1 score from 0.177 to 0.480—but this reliance on visual exemplars underscores the difficulty general-purpose models face in grounding subtle surgical concepts, such as anterior capsular radial folds, through text alone. Among medical-specific models, performance was highly polarized: QoQ-Med-VL demonstrated stable and competitive discrimination (F1 approximately 0.48), comparable to leading general-purpose models such as Qwen2.5-VL, whereas other adapters (eg, MedGemma and Lingshu) exhibited severe mode collapse. These findings indicate that domain pretraining alone does not guarantee fine-grained visual discrimination and may, in some architectures, introduce visual noise rather than clarity.

In contrast, automated task-specific AI model generators consistently produced supervised classifiers that approached the performance of junior glaucoma specialists, substantially narrowing, though not eliminating, the gap to expert observers. Human benchmarking further contextualized these results: general-purpose AI systems underperformed across all levels of clinical experience, remaining below even junior specialists despite few-shot prompting or fine-tuning, whereas task-specific models generated by an automated AI system achieved parity with junior specialists and medical trainees, although they did not reach the expert ceiling set by senior consultants.

Analysis of fine-tuning further highlighted the limitations of large general-purpose multimodal models for this task. Although fine-tuning Qwen2.5-VL improved sensitivity (recall from 0.555 to 0.603), it did so at the expense of specificity (precision from 0.804 to 0.684), yielding an overall F1 score (0.606) well below that of models generated by the automated task-specific approach. Collectively, these findings suggest that for specialized, high-stakes intraoperative visual recognition, task-specific supervised models generated by automated AI model generators currently offer a more viable near-term path to clinical utility than general-purpose multimodal reasoning.

### Clinical Implications

From a clinical and health system perspective, these findings suggest that deployment of surgical artificial intelligence should prioritize *task-specific validation* rather than reliance on general-purpose multimodal models. In cataract surgery, where brief failures to detect subtle intraoperative warning signs can lead to downstream complications, AI systems intended for intraoperative support must demonstrate reliability under the visual and temporal constraints of the operative task.

Automated generation of narrowly scoped, task-specific classifiers may therefore provide a safer and more practical entry point for surgical decision support, particularly for second-look warning systems or trainee supervision. These findings support the use of AI as a targeted adjunct during high-risk surgical phases rather than as a generalist replacement for surgeon judgment.

### Comparison With Prior Work

Recent ophthalmic AI research has shifted from general performance reporting toward specialized benchmarks that probe clinically meaningful reasoning. This trend is exemplified by text-based resources such as BELO, which evaluate the clinical knowledge and reasoning of large language models, as well as multimodal benchmarks including EyecareGPT and LMOD that focus on visual question answering.^29–31^ Datasets such as MM-Retinal-Reason have further attempted to bridge basic pattern recognition with higher-level clinical reasoning, and more recently, EH-Benchmark has extended evaluation to agent-driven workflows and hallucination-related failures.^25,32^

Despite these advances, standardized benchmarks for intraoperative surgical video interpretation remain limited. Prior work has largely focused on static images or outpatient diagnostic scenarios, whereas the present dataset targets dynamic surgical events and a subtle visual cue observable only within a narrow temporal window at capsular puncture. In addition, by benchmarking automated task-specific model generation alongside general-purpose AI systems, our study introduces a complementary methodological perspective: assessing whether task-specific supervised classifiers can be constructed without manual model engineering, which has received little attention in ophthalmic surgical AI.

### Limitations

Several limitations of our study warrant discussion. First, the dataset reflects the relatively low clinical prevalence of zonular instability, resulting in substantial class imbalance, with fold-negative cases constituting about 70% of cohorts. This imbalance likely biases general-purpose AI inference toward the majority class, favoring accuracy over sensitivity and contributing to the high precision but limited recall observed across models. To account for this imbalance and to ensure consistency between AI evaluation and human–AI comparison, we primarily reported macro-averaged precision, recall, and F1 scores, which weight both classes equally and provide a more appropriate assessment under skewed class distributions. Metrics such as sensitivity–specificity pairs or area under the receiver operating characteristic curve, while informative, were less suitable for direct comparison with human graders operating on the same imbalanced sample.

Second, the Singapore cohort was small, which limits the precision of external generalization estimates. Third, annotation of anterior capsular radial folds relied on binary labels without quantitative severity grading, and the frame-level annotations were performed by a single expert surgeon. Given the subtle and transient nature of early capsular folds, this approach introduces unavoidable subjectivity and precludes formal assessment of interrater reliability. Fourth, the frame sampling rate of 1 to 2 frames per second, although necessary for computational feasibility, may fail to capture brief visual cues occurring between sampled frames.

### Future Directions

Future research may address these limitations through enhanced annotation strategies and methodological refinement. Incorporation of multi-expert annotation would enable formal assessment of interrater reliability and support the development of continuous or ordinal severity scales that better capture the spectrum of zonular instability. From a video analysis perspective, adaptive keyframe selection strategies that estimate the distribution of visual content could help identify the most informative frames, capturing fleeting pathological manifestations while reducing redundant information.^33^ In addition, comprehensive assessment of intraoperative zonular instability may require integration of multiple correlated signs beyond the capsulorhexis phase, including equator visibility, capsulorhexis flutter, and postoperative decentration. Future work may therefore explore multitask learning approaches that jointly model these correlated indicators to improve overall diagnostic accuracy.

While this study focused on cataract surgery, the observed performance gap between general-purpose and task-specific AI systems is likely to be most pronounced in surgical contexts where diagnostic cues are brief, low-contrast, and temporally constrained. Similar characteristics are present in other procedures requiring fine-grained intraoperative judgment, such as capsule management, tissue plane identification, or early complication recognition.

## Conclusion

This multicenter diagnostic study demonstrates that general-purpose artificial intelligence systems do not reliably resolve brief, low-contrast cues in surgical video, even with few-shot visual examples, whereas automated task-specific AI model generators can produce supervised classifiers with substantially higher discriminative performance, approaching the level of junior glaucoma specialists. These findings highlight a methodological gap rather than a clinical determination and suggest that fine-grained surgical video interpretation is more likely to benefit from task-specific models generated by automated AI systems than from general-purpose AI. The dataset provides a reference point for future work on temporal grounding and surgical video analysis

## ARTICLE INFORMATION

**Author Contributions:** Y. Zhang and D. Liu had full access to all of the data in the study and take responsibility for the integrity of the data and the accuracy of the data analysis.

Concept and design: Y. Zhang, M. Tan, M. Wang, Y. Tham, C. Cheng, D. Liu.

Acquisition, analysis, or interpretation of data: Y. Zhang, L. Chen, W. Zhao, H Zhang, Chunyan Qiao, Zijian Liu, Charlotte Hui Chung.

Drafting of the manuscript: Y. Zhang, L. Chen, M. Tan, M. Wang, Y. Tham, C. Cheng, D. Liu.

Critical review of the manuscript for important intellectual content: Y. Zhang, L. Chen, M. Tan, M. Wang, Y. Tham, C. Cheng, D. Liu.

Statistical analysis: Y. Zhang, L. Chen.

Administrative, technical, or material support: M. Tan, M. Wang, Y. Tham, C. Cheng, D. Liu.

### Data and Sharing Statement

**Data Sharing:** Deidentified full-length surgical videos supporting the findings of this study will be made publicly available. The released dataset includes complete phacoemulsification videos from the Beijing Tongren Hospital cohort, with **clip-level annotations** indicating major surgical phases (eg, incision, continuous curvilinear capsulorhexis, hydrodissection, phacoemulsification, cortical aspiration, intraocular lens implantation) and multiple intraoperative signs related to zonular instability. These annotations include equator visibility, anterior capsular radial folds at capsulorhexis initiation, capsulorhexis flutter during cortical aspiration, and capsulorhexis deviation after cortical aspiration. The dataset is available at https://huggingface.co/datasets/leonChen/Tongren-Zonular-Video. A data dictionary and detailed annotation descriptions are provided within the repository.

**Code:** All source code used for model training, evaluation, and analysis is publicly available at https://github.com/cherishleon/ZonularBench.

**Timing and Access:** The data and code are available at the time of publication and will remain accessible without time restriction.

**Conditions of Use:** The data may be used for noncommercial research purposes, including method development, benchmarking, and secondary analyses related to surgical video analysis. Users are required to cite the present study and comply with the repository’s license terms.

**Additional Information:** No individual patient identifiers are included in the shared data. Institutional review board approval was obtained at the contributing institutions, and data sharing complies with applicable ethical and privacy regulations.

## Data Availability

The dataset is available at https://huggingface.co/datasets/leonChen/Tongren-Zonular-Video.

## Supplementary Material

### A. Overview of the Vision-Language Models inference pipelines

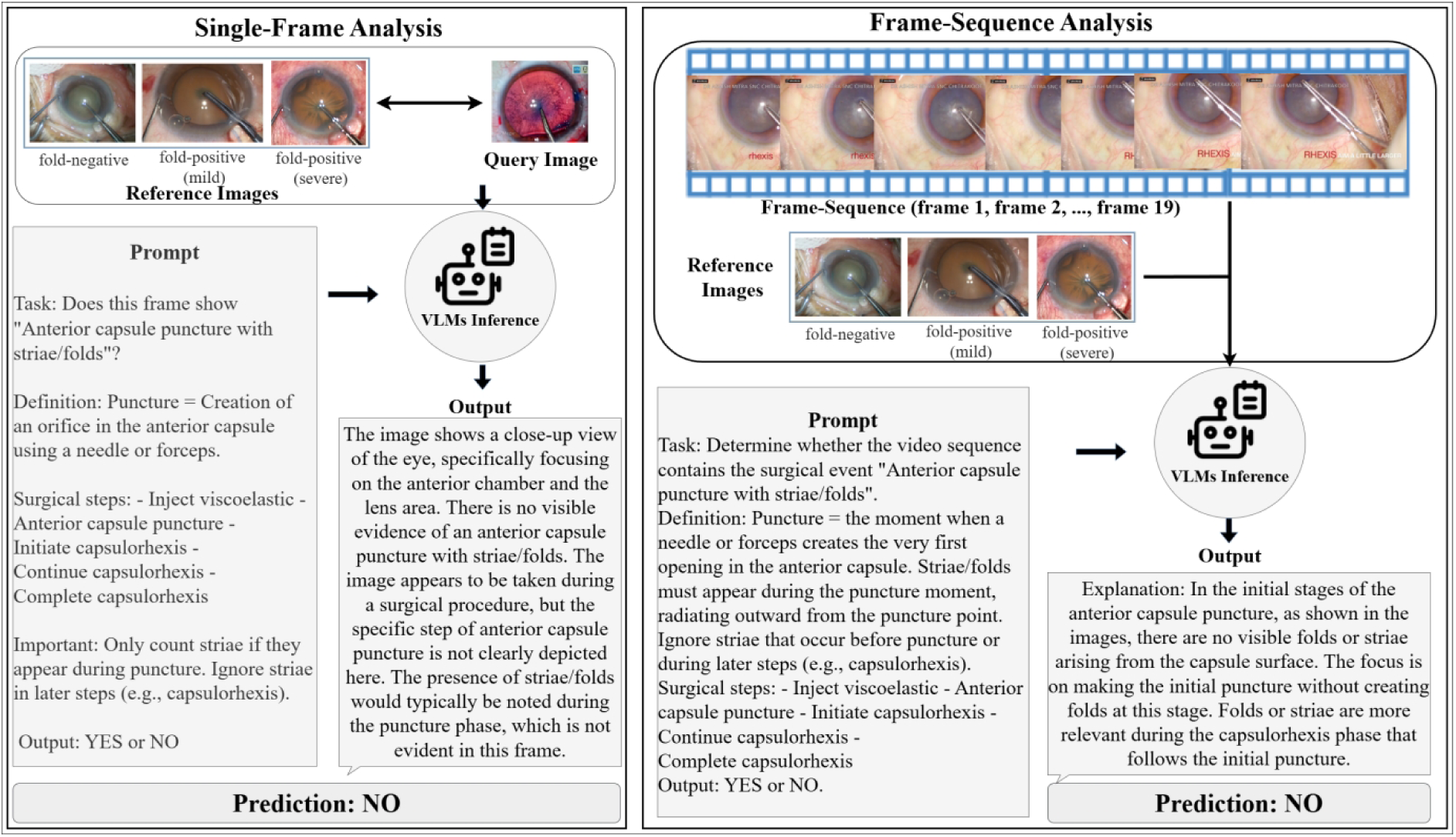

#### Overview of the Vision-Language Models inference pipelines

The top panel shows the Single-Frame Analysis, where reference images, a query image, and a task definition prompt are processed by the Vision-Language Model to generate a prediction. The bottom panel shows the Frame-Sequence Analysis, utilizing a sequence of frames to provide temporal context, along with reference images and a specific prompt, leading to a prediction.

### B. Prompt for Vision-Language Models for Single-Frame Analysis

**V1:** ’’’[MEDICAL DOCUMENT - CATARACT SURGERY ANALYSIS]

You are analyzing medical/surgical footage from a cataract extraction procedure (phacoemulsification). This is a routine ophthalmic surgical procedure.

Task: Does this frame show “Anterior capsule puncture with striae/folds”?

Definition: Puncture = Creation of an orifice in the anterior capsule using a needle or forceps. Surgical steps: - Inject viscoelastic - Anterior capsule puncture - Initiate capsulorhexis - Continue capsulorhexis - Complete capsulorhexis

Important: Only count striae if they appear during puncture. Ignore striae in later steps (e.g., capsulorhexis).

Output: YES or NO ’’’

**V2:** ’’’[MEDICAL DOCUMENT - OPHTHALMIC SURGICAL ANALYSIS]

You are reviewing frames from a cataract extraction procedure. This is a standard medical/surgical documentation task.

Question: Does this image show “Anterior capsule puncture with striae”? Criteria:

- Look for a puncture being made with a needle/forceps.

- If folds radiate from this puncture point, answer YES.

- If no folds, or folds appear later in capsulorhexis, answer NO. Output: YES or NO. ’’’

**V3:** ’’’[MEDICAL DOCUMENT - OPHTHALMIC SURGICAL ANALYSIS]

You are reviewing frames from a cataract extraction procedure. This is a standard medical/surgical documentation task.

Instruction: Assess whether this frame shows “Anterior capsule puncture with striae”.

Checkpoints: Puncture = a needle or forceps is creating the first opening in the anterior capsule. Striae/folds must radiate from this puncture point at that moment. If striae appear only later in capsulorhexis, do not count.

Answer with: YES or NO. ’’’

### C. Prompt for Vision-Language Models for Frame-Sequence Analysis

**V1:** ’’’Task: Determine whether the video sequence (seq) contains the surgical event “Anterior capsule puncture with striae/folds”.

Definition:

- Puncture = the moment when a needle or forceps creates the very first opening in the anterior capsule.

- Striae/folds must appear *during the puncture moment*, radiating outward from the puncture point.

- Ignore striae that occur before puncture or during later steps (e.g., capsulorhexis).

Surgical steps: - Inject viscoelastic - Anterior capsule puncture - Initiate capsulorhexis - Continue capsulorhexis - Complete capsulorhexis

Output: YES or NO.’’’

**V2:** ’’’Question: During the anterior capsule puncture process shown in this video sequence, do striae/folds appear?

Criteria:

- Locate the moment when the needle/forceps initiates the anterior capsule puncture.

- Answer YES if folds/striae arise from the capsule surface at that moment.

- Answer NO if: puncture happens without folds, or folds occur only before or after the puncture period.

Important:

Evaluate only the puncture phase, not later capsulorhexis. Output: YES or NO.’’’

**V3:** ’’’Instruction: Inspect the full video sequence and determine whether striae/folds appear during the anterior capsule puncture.

Checkpoints:

- Identify the puncture moment when the needle or forceps creates the initial opening.

- Confirm whether striae/folds appear at that same time.

- Ignore folds that occur outside the puncture process. Answer with: YES or NO.’’’

### D. Prompt for Automated AI model generators

’’’*You are an advanced code generation and optimization agent. Your task is to automatically build, train, and evaluate a binary classification model for surgical image classification using PyTorch and CUDA device 0*.

*Project structure: The current directory contains three CSV files:*

● *train_data.csv — training set*
● *val_data.csv — validation set*
● *test_data.csv — test set*

*Each CSV file has two columns:*

● *image_path — the file path to each image*
● *abnorm — binary label (0 = normal, 1 = abnormal)*

*Your goals: Automatically generate complete runnable code that:*

*1. Loads and preprocesses the dataset from the CSVs (train/val/test)*
*2. Uses appropriate image transformations (resize, normalization, augmentation for train)*
*3. Creates PyTorch Dataset and DataLoader classes*
*4. Builds an efficient CNN or pretrained backbone (e.g. ResNet50, EfficientNet, or similar) for binary classification*
*5. Supports GPU acceleration with cuda:0*
*6. Periodically evaluates on the validation set during training and prints accuracy, precision, recall, F1, AUROC*
*7. Saves the best model checkpoint based on validation F1*
*8. After training, runs final evaluation on the test set and prints a classification report*
*9. Include all necessary imports and ensure reproducibility (set seeds)*
*10. Make the code modular and readable, with functions for dataset loading, model definition, training loop, and evaluation*
*11. Automatically optimize model hyperparameters (e.g., learning rate, batch size, number of layers, dropout) if possible*
*12. At the end, print total training time and final metrics on the test set*

*Environment:PyTorch≥2.0,CUDAdevice0.Usemixed-precisiontraining ifbeneficial(torch.amp).*

*After completing the code, you need to run it, automatically analyze the results, and then optimize the code.*’’’

### E. Prompt for LLM-as-Judge

’’’ You are an expert code reviewer tasked with evaluating the quality of automatically generated Python code for a deep learning image classification project. Please assess the provided code across five dimensions, assigning a score from 0 to 20 points for each dimension based on the criteria below. Provide a score and brief justification for each dimension.

#### Evaluation Criteria

1. Code Organization and Modularity (0-20 points)

● Logical code structure with clear separation of concerns (data loading, model definition, training, evaluation)
● Modular design with reusable functions and classes
● Well-defined abstraction layers
● Avoidance of code duplication

#### Scoring Guide

● 18-20: Excellent organization with highly modular, reusable components and clear separation of concerns
● 14-17: Good organization with reasonable modularity, minor improvements possible
● 10-13: Adequate organization but lacks clear modularity or has some structural issues
● 6-9: Poor organization with significant structural problems or code duplication
● 0-5: Minimal organization, monolithic code structure

2. Documentation and Readability (0-20 points)
  ● Clear and meaningful variable and function naming conventions
  ● Consistent code style and formatting
  ● Readable code flow and logic

#### Scoring Guide

● 18-20: Excellent readability with intuitive naming and consistent style throughout
● 14-17: Good readability with clear naming, minor inconsistencies
● 10-13: Adequate readability but some confusing names or style inconsistencies
● 6-9: Poor readability with unclear naming or inconsistent formatting
● 0-5: Very poor readability, cryptic naming, or chaotic formatting

3. Error Handling and Robustness (0-20 points)
  ● Exception handling mechanisms for critical operations (file I/O, model loading, GPU operations)
  ● Input validation and boundary condition checks
  ● Graceful handling of missing or corrupted data
  ● Fault tolerance in data loading and training pipelines

#### Scoring Guide

● 18-20: Comprehensive error handling with robust exception management and validation
● 14-17: Good error handling for most critical operations
● 10-13: Basic error handling present but incomplete coverage
● 6-9: Minimal error handling, vulnerable to common failure models
● 0-5: No error handling or verification mechanisms

4. Software Engineering Best Practices (0-20 points)
  ● Reproducibility through proper random seed configuration
  ● Standardized data loading workflows
  ● Configuration management (avoiding hard-coded values, using arguments or config files)
  ● Comprehensive logging and experiment tracking
  ● Result visualization capabilities (training curves, confusion matrices, etc.)
  ● Model checkpoint management with ability to resume training

#### Scoring Guide

● 18-20: Exemplary implementation of all best practices
● 14-17: Most best practices implemented, minor gaps
● 10-13: Some best practices present but significant omissions
● 6-9: Few best practices implemented, major gaps
● 0-5: Minimal or no adherence to best practices

5. Code Efficiency (0-20 points)
  ● Efficient data loading pipelines (multi-worker data loaders, prefetching, caching)
  ● GPU utilization and parallel processing optimizations
  ● Memory usage optimization (batch size management, gradient accumulation if needed)
  ● Implementation of performance-enhancing features (mixed-precision training, efficient transforms)

#### Scoring Guide

● 18-20: Highly optimized with multiple efficiency techniques implemented
● 14-17: Well-optimized with most efficiency features present
● 10-13: Adequate efficiency with basic optimizations
● 6-9: Suboptimal efficiency, missing key optimizations
● 0-5: Inefficient implementation with no optimization considerations

### F. The specific performance of Vision-Language Models at the frame-level, clip-level and frame-sequence across three prompts

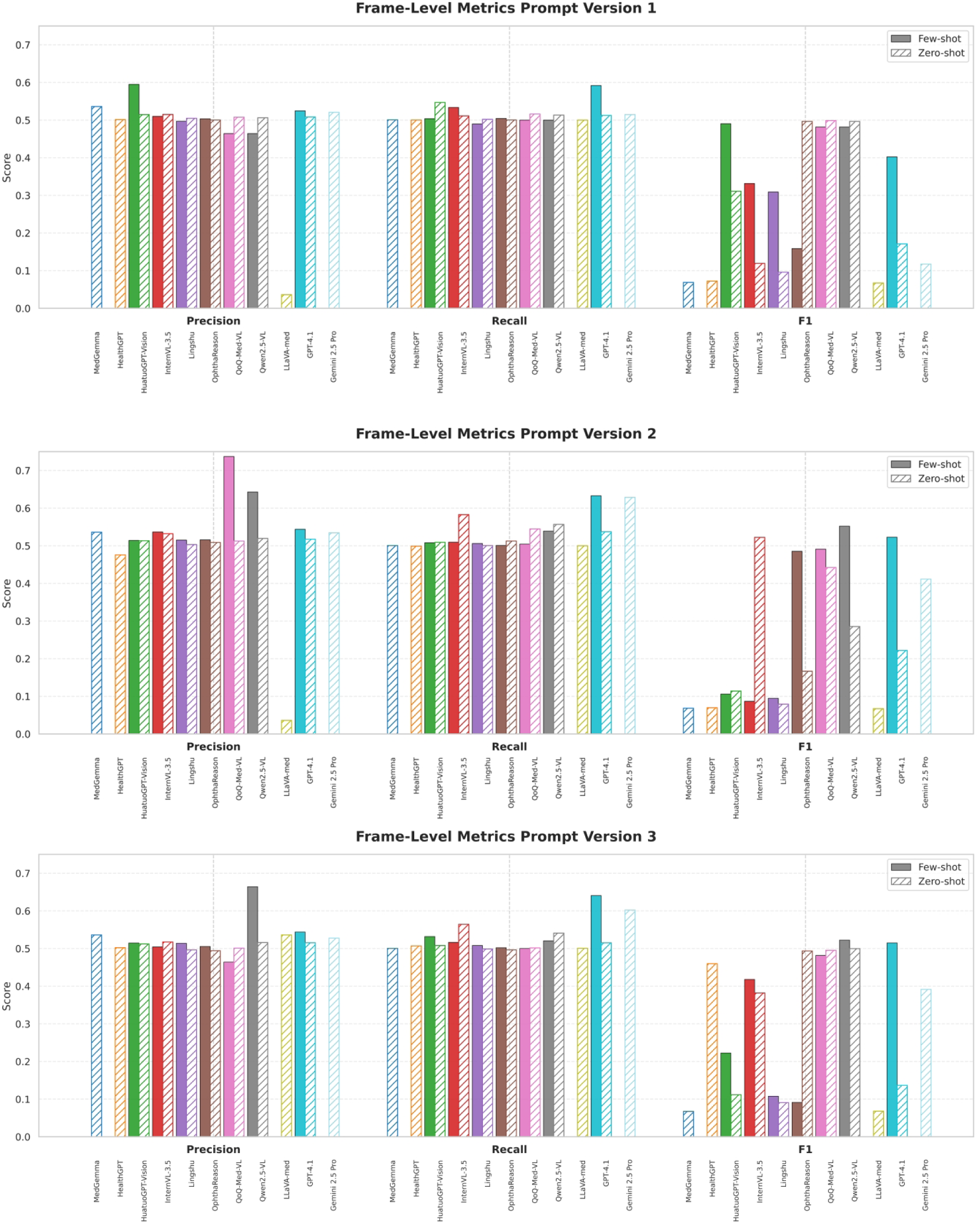

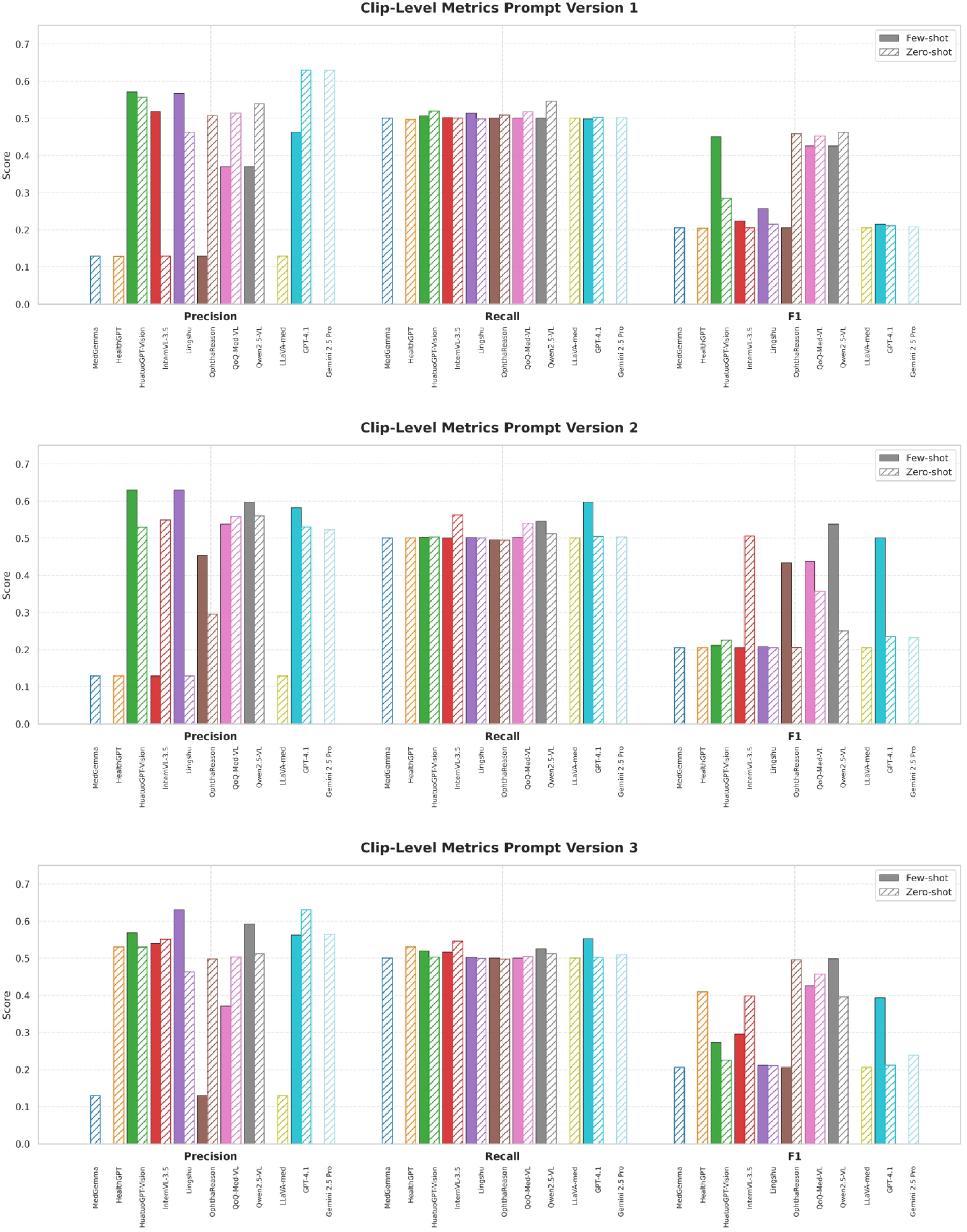

**Table G-1.** Comprehensive Performance Benchmark of Vision-Language Models across Multi-Granularity Analysis Tasks. This Table provides a general benchmark of Vision-Language Models, detailing their performance on the full test set across three tasks: Frame-Level (top 3 models), Clip-Level (top 3 models), and Frame-Sequence analysis (all evaluated models). Data are presented as Mean ± Standard Deviation for each metric and setting. The result is macro-averaged.

|  |  |  |  |  |  |
| --- | --- | --- | --- | --- | --- |
| **1. Frame-Level Analysis** | Qwen2.5-VL | Few-shot | $0.590 \pm 0.090$ | $0.520 \pm 0.016$ | $0.519 \pm 0.029$ |
| | QoQ-Med-VL | Few-shot | $0.555 \pm 0.129$ | $0.501 \pm 0.002$ | $0.485 \pm 0.004$ |
| | GPT-4.1 | Few-shot | $0.537 \pm 0.009$ | $0.622 \pm 0.021$ | $0.480 \pm 0.055$ |
| **2. Clip-Level Analysis** | Qwen2.5-VL | Few-shot | $0.520 \pm 0.106$ | $0.524 \pm 0.019$ | $0.487 \pm 0.046$ |
| | QoQ-Med-VL | Few-shot | $0.426 \pm 0.079$ | $0.501 \pm 0.001$ | $0.430 \pm 0.006$ |
| | QoQ-Med-VL | Zero-shot | $0.526 \pm 0.024$ | $0.521 \pm 0.015$ | $0.422 \pm 0.046$ |
| **3. Frame-Sequence Analysis** | Qwen2.5-VL | Few-shot | $0.589 \pm 0.256$ | $0.509 \pm 0.012$ | $0.461 \pm 0.054$ |
| | QoQ-Med-VL | Few-shot | $0.426 \pm 0.097$ | $0.501 \pm 0.002$ | $0.432 \pm 0.011$ |
| | InternVL-3.5 | Few-shot | $0.425 \pm 0.034$ | $0.437 \pm 0.025$ | $0.320 \pm 0.081$ |
| | HuatuoGPT-Vision | Few-shot | $0.571 \pm 0.015$ | $0.519 \pm 0.002$ | $0.272 \pm 0.013$ |
| | Lingshu | Few-shot | $0.433 \pm 0.267$ | $0.506 \pm 0.009$ | $0.237 \pm 0.050$ |

### G. Result for Automatic AI model generators

**Table G-1.** Automatic AI model generators performance on the test set.

| Generators | Precision | Recall | F1 | AUC |
| --- | --- | --- | --- | --- |
| Copilot | 0.842 | **0.903** | **0.870** | 0.959 |
| Qoder | **0.851** | 0.866 | 0.859 | **0.960** |
| CodeX | 0.840 | 0.862 | 0.850 | 0.954 |
| AIDE | 0.470 | 0.500 | 0.484 | 0.500 |

**Table G-2.** Code quality (LLM-as-judge, 0–20).

| Generators | Organization & Modularity | Documentation & Readability | Error Handling & Robustness | SE Best Practices | Code Efficiency |
| --- | --- | --- | --- | --- | --- |
| Copilot | **19** | **18** | **17** | **19** | **18** |
| CodeX | **19** | 17 | 15 | 18 | **19** |
| Qoder | 17 | 16 | 15 | 17 | **18** |
| AIDE | 10 | 12 | **5** | 8 | 11 |

**Figure G.**
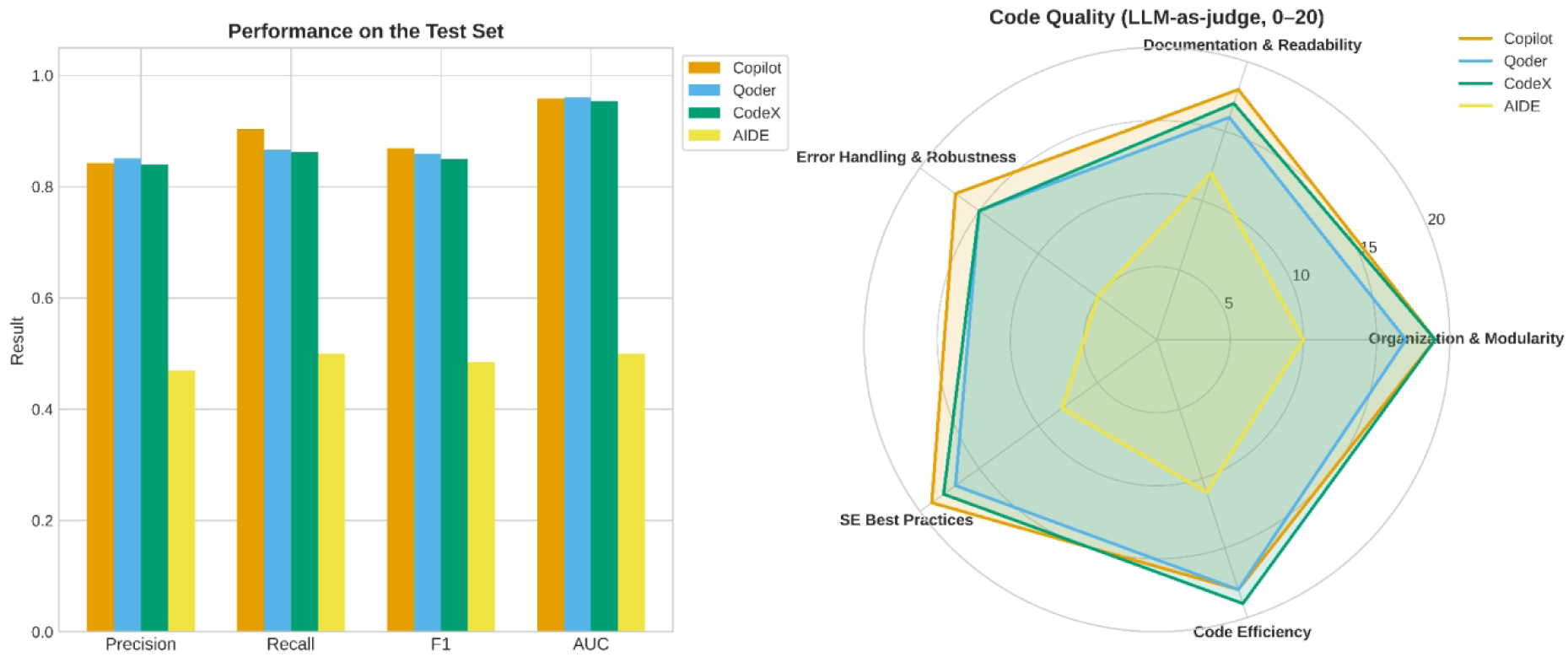
Comparative Performance and Code Quality of Four Automatic AI model generators. The bar chart (left) displays classification metrics on the test set, while the radar chart (right) illustrates code quality scores (0–20) assessed by five dimensions.

